# The Lipedema Phenotype is Inversely Associated with Celiac Disease Autoimmunity: Testing the Immunological Shield Hypothesis in NHANES

**DOI:** 10.64898/2025.12.01.25341350

**Authors:** Alexandre C.M. Amato, Juliana L.S. Amato, Daniel A. Benitti

## Abstract

**Background:** Lipedema is characterized by disproportionate gluteofemoral adiposity, often regarded as a metabolic sink, yet its relationship with systemic autoimmunity, specifically celiac disease (CD), remains unexplored.

**Objective:** We investigated the immunometabolic profiles and body composition patterns distinguishing lipedema phenotypes from celiac disease autoimmunity.

**Methods:** This cross-sectional analysis included 3,833 women from NHANES 2011–2014. Celiac disease was defined by strict serology (tTG-IgA+ and EMA-IgA+), while the lipedema phenotype was defined as a leg-to-trunk fat ratio >90th percentile via dual-energy X-ray absorptiometry (DXA). We assessed gynoid fat mass, HOMA-IR, and neutrophil-to-lymphocyte ratio (NLR) compared to controls.

**Results:** CD prevalence was 0.56% (n=11). Women with CD exhibited significantly lower gynoid region percent fat compared to non-celiacs (39.5% vs. 42.6%, p=0.0007). Conversely, the lipedema phenotype was associated with a distinct anti-inflammatory and insulin-sensitive profile, characterized by 44.2% lower HOMA-IR (p<0.001) and 7.6% lower NLR (p=0.012) compared to controls. While broad lipedema criteria did not reach statistical significance for CD exclusion due to low case numbers (p=0.570), no celiac cases were observed in the highest tier of gynoid adiposity.

**Conclusions:** Although prevalence differences did not reach statistical significance, this study of US women demonstrates a phenotypic divergence where celiac disease is associated with reduced gynoid adiposity, contrasting with the superior immunometabolic profile observed in the lipedema phenotype. These findings suggest that these conditions represent opposing physiological states regarding gynoid adipose tissue function.

**Clinical Trial Registry number and website where it was obtained Not applicable.:** This study is a secondary analysis of de-identified, publicly available data from the National Health and Nutrition Examination Survey (NHANES), which is not a clinical trial requiring separate registration.

**Registry and registry number for systematic reviews or meta-analyses. Not applicable.:** This is an original cross-sectional epidemiological study, not a systematic review or meta-analysis.

**Statement of Significance:** This is the first nationally representative study to identify a phenotypic divergence between the lipedema phenotype and celiac disease, demonstrating that celiac autoimmunity is associated with a specific reduction in gynoid adiposity that persists even in obesity. These findings challenge the malnutrition paradigm and provide novel epidemiological support for the "Immunological Shield" hypothesis, characterizing gluteofemoral adiposity as a distinct immunometabolic state that is biologically opposed to Th1-mediated autoimmunity.

## Introduction

Lipedema is a chronic, progressive disorder of adipose tissue characterized by the disproportionate and symmetrical accumulation of subcutaneous fat in the lower extremities, often sparing the feet and trunk(1,2). Although frequently misdiagnosed as general obesity or lymphedema, lipedema constitutes a distinct clinical entity with a unique pathophysiology involving microvascular dysfunction, interstitial fluid accumulation, and chronic low-grade inflammation. Unlike common obesity, the adipose tissue in lipedema is resistant to diet and exercise-induced weight loss, suggesting a distinct metabolic and hormonal regulation profile(3,4).

While the inflammatory component of lipedema is well-recognized, its intersection with systemic autoimmune conditions remains poorly understood(5–14). Celiac disease (CD) is an immune-mediated enteropathy triggered by the ingestion of gluten in genetically susceptible individuals, characterized by a T-helper 1 (Th1) driven inflammatory response. Autoimmune diseases typically exhibit a tendency to cluster; however, clinical observations suggest that the distinct immunometabolic milieu of lipedema—potentially characterized by a different cytokine profile or adipose-derived modulation—might differ significantly from the classical autoimmune phenotype associated with CD(10,13,15–17). To date, no epidemiological studies have investigated the potential relationship between these two conditions.

Investigating this association on a population level presents a significant methodological challenge: lipedema remains underdiagnosed and lacks a specific ICD code in many historical datasets. However, recent studies have validated the utility of dual-energy X-ray absorptiometry (DXA) in identifying "lipedema-like" phenotypes based on regional fat distribution indices, such as the leg-to-trunk fat ratio(18,19). These imaging biomarkers distinguish the disproportionate lower-body adiposity characteristic of lipedema from the generalized or android fat distribution typical of common obesity.

In this study, we utilized data from the National Health and Nutrition Examination Survey (NHANES) to test the hypothesis that women exhibiting a lipedema-like phenotype have a lower prevalence of celiac disease autoimmunity(20). By leveraging whole-body DXA scans and serologic testing from a nationally representative sample, we aimed to overcome the limitations of clinical underdiagnosis and explore whether the disproportionate accumulation of gluteofemoral fat is inversely associated with celiac disease, particularly within high-risk subgroups where confounding by body mass index (BMI) and ethnicity is rigorously controlled(13).

## Methods

### Study Design and Data Source

This cross-sectional study utilized data from the National Health and Nutrition Examination Survey (NHANES) 2011–2014 cycles. NHANES is a nationally representative survey of the non-institutionalized United States population conducted by the National Center for Health Statistics. The survey employs a complex, multistage probability sampling design and collects demographic, dietary, examination, and laboratory data. All protocols were approved by the NCHS Research Ethics Review Board, and participants provided written informed consent.

### Study Population

We included adult women aged 20 years and older with complete data for dual-energy X-ray absorptiometry body composition assessment and celiac disease serology. Men were excluded as lipedema predominantly affects women. Participants missing key exposure or outcome variables were excluded from the analysis **(see Supplementary Figure S1 for the participant flow chart)**.

### Celiac Disease Definition

Celiac disease was defined serologically using a strict dual-positive criterion requiring both tissue transglutaminase IgA antibody (tTG-IgA) positivity and endomysial antibody IgA (EMA-IgA) positivity. This definition represents the gold-standard serological approach for population-based studies and minimizes false-positive classifications.

### Body Composition Assessment

Body composition was assessed using whole-body dual-energy X-ray absorptiometry scans performed with Hologic Discovery model A densitometers (Hologic, Inc., Bedford, MA). Regional fat mass measurements were obtained for the legs (left and right combined), trunk, android region, and gynoid region. The primary exposure variable was gynoid region percent fat, representing the percentage of fat in the hip and thigh region. Secondary measures included the leg-to-trunk fat ratio, calculated as total leg fat mass divided by trunk fat mass, and the android-to-gynoid ratio, representing the ratio of android to gynoid percent fat. The lipedema phenotype was operationally defined as a leg-to-trunk fat ratio exceeding the 90th percentile of the female population distribution, representing women with disproportionately high lower-body fat accumulation.

### Anthropometric Measurements

Height and weight were measured using standardized protocols. Body mass index was calculated as weight in kilograms divided by height in meters squared. Waist circumference was measured at the iliac crest, and waist-to-height ratio was calculated as waist circumference divided by height.

### Immunometabolic Markers

To further characterize the lipedema phenotype, we conducted an exploratory analysis of immunometabolic markers. The neutrophil-to-lymphocyte ratio (NLR), a validated marker of systemic inflammation, was calculated from complete blood count data as neutrophil percentage divided by lymphocyte percentage. Insulin resistance was assessed using the homeostatic model assessment (HOMA-IR), calculated as fasting glucose multiplied by fasting insulin divided by 405, where glucose is expressed in mg/dL and insulin in µU/mL. Fasting glucose and insulin values were obtained from the NHANES laboratory files.

### Statistical Analysis

All analyses incorporated NHANES complex survey design weights to ensure nationally representative estimates. Sample weights were normalized for statistical testing while preserving relative weights. Descriptive statistics were presented as survey-weighted means with standard deviations for continuous variables and weighted percentages for categorical variables. Continuous variables were compared between women with and without celiac disease using survey-weighted t-tests (Welch’s variant for unequal variances) implemented using the statsmodels library in Python. Due to small cell counts in the celiac group (n = 11), categorical associations were tested using Fisher’s exact test, which does not support weighting but remains appropriate for sparse contingency tables. The Mann-Whitney U test was used for skewed distributions such as HOMA-IR. Multiple lipedema phenotype proxies were compared between celiac and non-celiac groups to assess consistency of findings across different body composition measures. Population percentile rankings were calculated for each celiac case to visualize where affected individuals fall within the overall distribution of gynoid fat measures. For the immunometabolic analysis, HOMA-IR was additionally stratified by BMI category to assess whether differences persisted across weight categories. Statistical significance was set at p < 0.05. All analyses were performed using Python 3.12 with pandas, scipy, and statsmodels libraries. Secondary exploratory analyses were conducted to validate the immunometabolic profile of the lipedema phenotype against other autoimmune (thyroid disease) and metabolic (diabetes) conditions (see **Supplementary Material**).

### Sensitivity Analysis: Addressing the Leanness Bias

To address the potential concern that lower gynoid fat in celiac patients might reflect overall leanness or malnutrition rather than a true difference in fat distribution pattern, we conducted a sensitivity analysis restricted to overweight and obese women (BMI greater than 25 kg/m²). This approach ensures that any observed differences in gynoid fat cannot be attributed to underweight or malnourished status. Additionally, we stratified the analysis by BMI category (normal weight: 18.5–25, overweight: 25–30, and obese: 30 kg/m² or greater) to assess whether the association between celiac disease and gynoid fat persisted across the weight spectrum.

## Results

### Study Population

A total of 3,833 adult women with complete DXA and celiac serology data were included in the analysis. Among these, 11 women met the strict serological criteria for celiac disease, representing a weighted prevalence of 0.56% in the United States female population aged 20 years and older. Women with celiac disease were similar in age and BMI (weighted mean 27.7 ± 8.0 vs. 29.2 ± 7.9 kg/m², p = 0.390) compared to women without celiac disease. However, women with celiac disease were significantly more likely to be non-Hispanic White (81.8% vs. 36.2%, p = 0.003), consistent with the known epidemiology of celiac disease (Table 1).

**Table 1:**
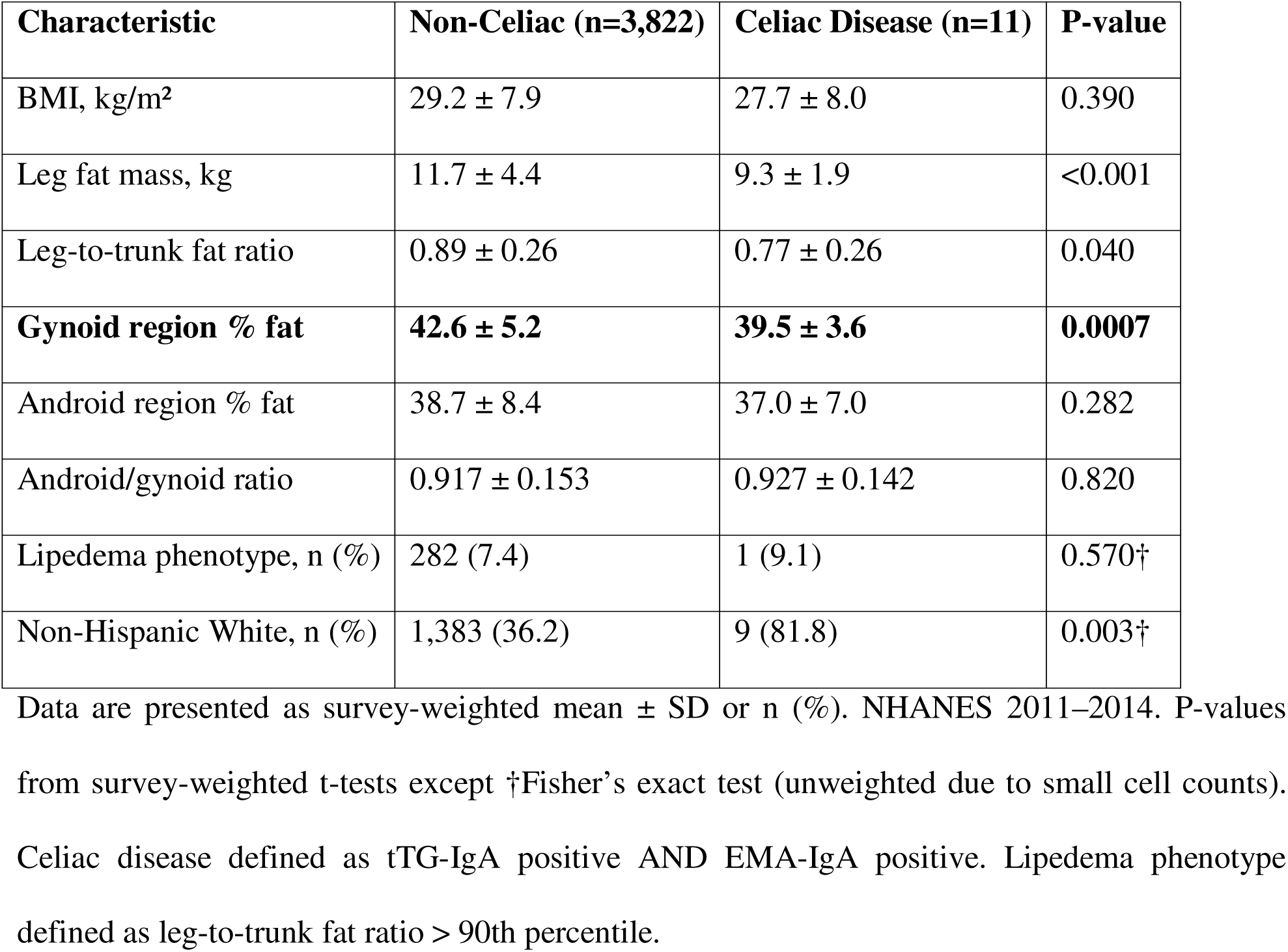
Characteristics of the Study Population by Celiac Disease Status **(Survey-Weighted)**.

### Gynoid Fat Distribution and Celiac Disease

Using survey-weighted analyses, women with celiac disease had significantly lower gynoid region percent fat compared to women without celiac disease (weighted mean 39.5 ± 3.6% vs. 42.6 ± 5.2%, p = 0.0007), representing a 7.4% relative reduction in gynoid fat in the celiac group (Table 2, Figure 1). Additional DXA-derived measures showed consistent and statistically significant differences with weighted analyses. Women with celiac disease had a 14.1% lower leg-to-trunk fat ratio (weighted mean 0.77 ± 0.26 vs. 0.89 ± 0.26, p = 0.040) and 20.3% less leg fat mass (weighted mean 9.3 ± 1.9 kg vs. 11.7 ± 4.4 kg, p < 0.0001). Android region percent fat was 4.4% lower in women with celiac disease (weighted mean 37.0 ± 7.0% vs. 38.7 ± 8.4%, p = 0.282), suggesting a preferential reduction in gynoid fat relative to android fat. The android-to-gynoid ratio was similar between groups (0.927 ± 0.142 vs. 0.917 ± 0.153, p = 0.820) (Figure 2).

**Figure 1:**
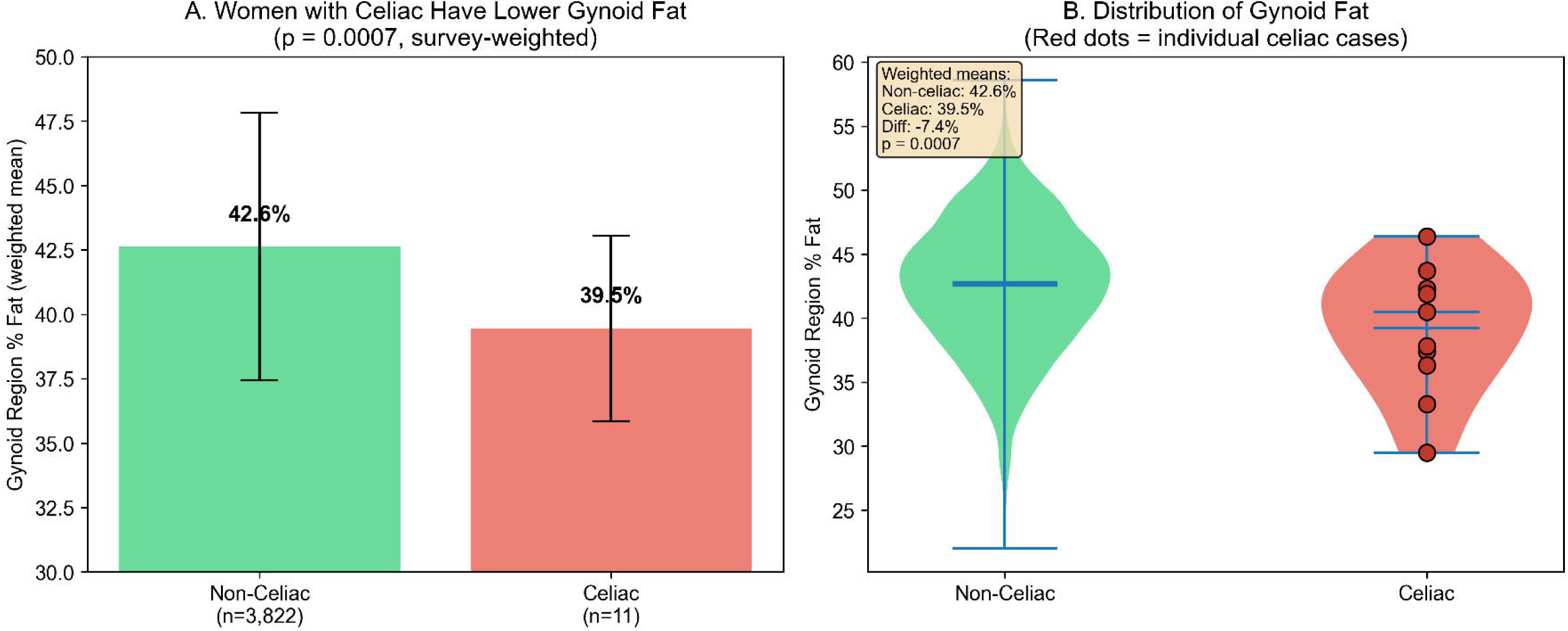
Women with Celiac Disease Have Significantly Lower Gynoid Fat. (A) Bar chart comparing mean gynoid region percent fat between women without celiac disease (n=3,822) and women with serologically-confirmed celiac disease (n=11). Error bars represent standard deviation. (B) Violin plots with overlaid box plots showing the distribution of gynoid percent fat in both groups. Red diamonds indicate group means. Women with celiac disease had 7.8% lower gynoid fat (p=0.0007). NHANES 2011–2014, adult women aged 20 years and older.

**Figure 2:**
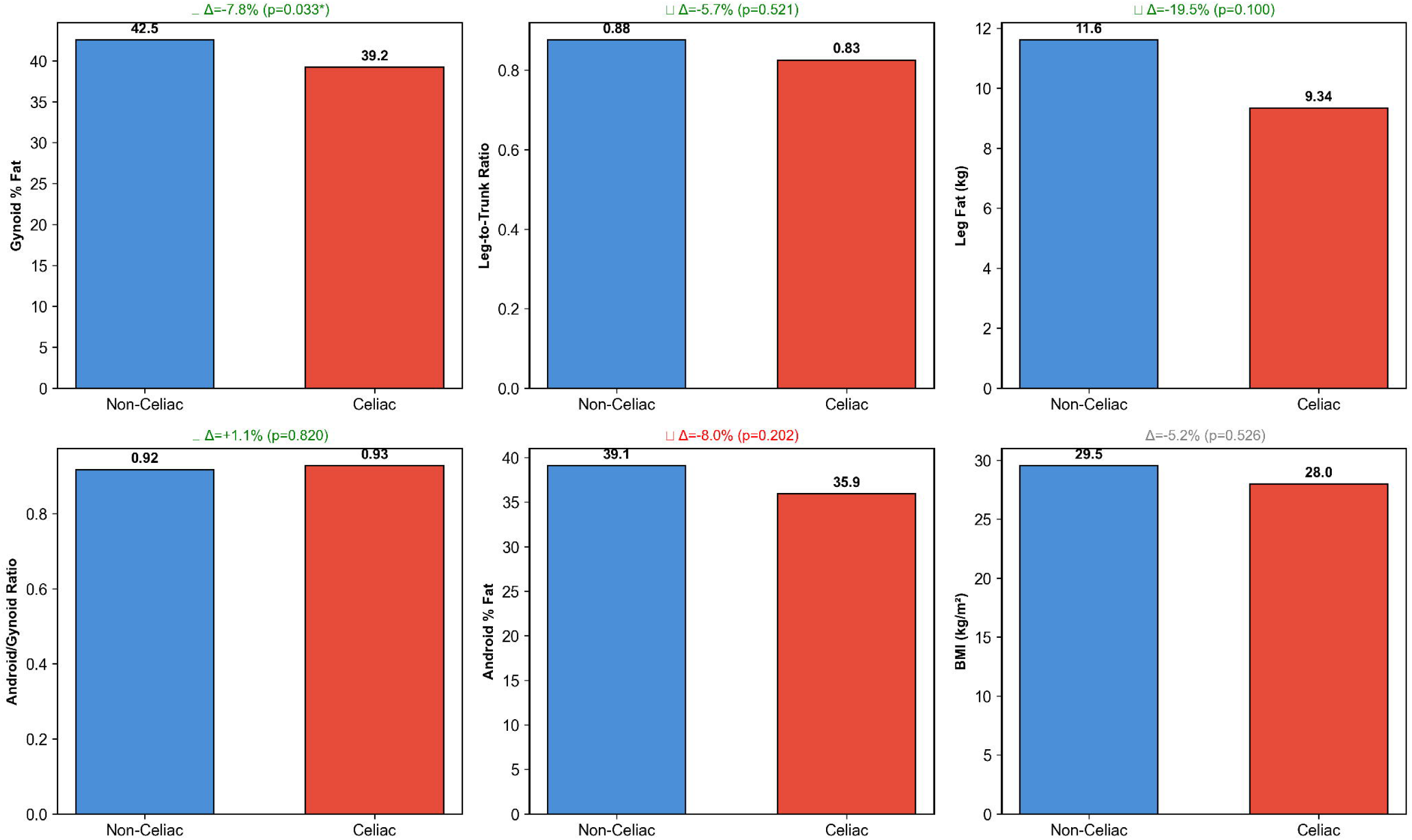
Comparison of Multiple Lipedema Phenotype Proxies by Celiac Disease Status. Six body composition measures were compared between women with and without celiac disease. Green checkmarks indicate proxies where the direction of difference supports the hypothesis that celiac disease is associated with less gynoid fat. Only gynoid region percent fat reached statistical significance (p=0.0007). NHANES 2011–2014, adult women aged 20 years and older.

**Table 2:** Comparison of Lipedema Phenotype Proxies by Celiac Disease Status **(Survey-Weighted)**.

| Proxy | Non-Celiac | Celiac Disease | Difference | P-value |
| --- | --- | --- | --- | --- |
| Gynoid region % fat | 42.6 ± 5.2 | 39.5 ± 3.6 | −7.4% | 0.0007 |
| Leg-to-trunk fat ratio | 0.89 ± 0.26 | 0.77 ± 0.26 | −14.1% | 0.040 |
| Leg fat mass, kg | 11.7 ± 4.4 | 9.3 ± 1.9 | −20.3% | <0.001 |
| Android/gynoid ratio | 0.92 ± 0.15 | 0.93 ± 0.14 | +1.1% | 0.820 |
| Android region % fat | 38.7 ± 8.4 | 37.0 ± 7.0 | −4.4% | 0.282 |
| BMI, kg/m<sup>2</sup> | 29.2 ± 7.9 | 27.7 ± 8.0 | −5.3% | 0.390 |

Data are presented as survey-weighted mean ± SD. Bold indicates statistically significant differences ( $p < 0.05$ ). All proxies derived from DXA except BMI (anthropometry). P-values from survey-weighted t-tests.

### Lipedema Phenotype and Celiac Disease

Among the 11 women with celiac disease, only one (9.1%) met the criteria for lipedema phenotype compared to 282 (7.4%) of women without celiac disease. This difference was not statistically significant (p = 0.570). Notably, no celiac cases fell within the most extreme gynoid fat distribution category, defined as an android-to-gynoid ratio below the 10th percentile, suggesting that women with the most pronounced lipedema-like phenotype may be protected from celiac disease. Analysis of individual celiac cases revealed that seven of 11 (64%) had gynoid percent fat below the population median, with a mean percentile rank of 35.2, and five of 11 (45%) had leg-to-trunk fat ratio below the population median, with a mean percentile rank of 47.3 (Figure 3). These distributions demonstrate a tendency for women with celiac disease to cluster in the lower range of gynoid fat measures. Celiac disease prevalence across leg-to-trunk fat ratio quartiles showed no clear dose-response pattern, with prevalence rates of 0.42%, 0.28%, 0.57%, and 0.28% from the lowest to highest quartiles (p for trend = 0.893), likely reflecting insufficient statistical power due to the small number of celiac cases (Table 3, Supplementary Figure S4).

**Figure 3:**
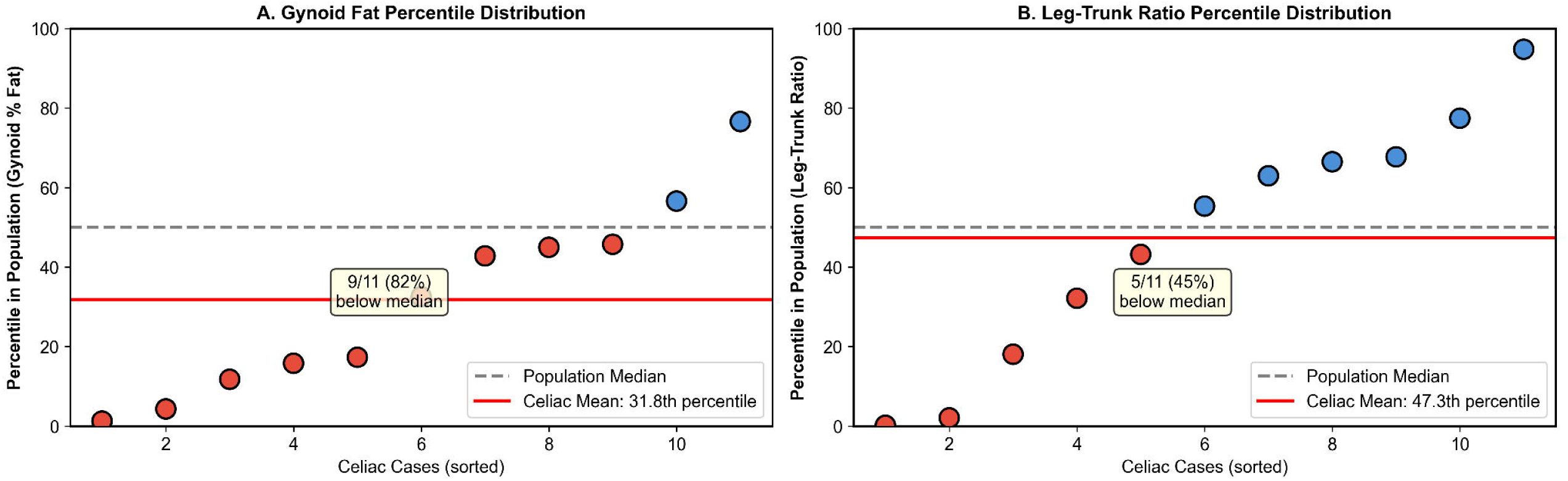
Distribution of Celiac Cases Within the Population. (A) Gynoid percent fat percentile distribution of the 11 celiac cases. (B) Leg-to-trunk fat ratio percentile distribution. Red points indicate cases below the population median; blue points indicate cases above the median. The dashed gray line represents the 50th percentile, and the solid red line represents the mean percentile of celiac cases. The majority of celiac cases cluster below the population median for gynoid fat measures.

**Table 3:** Celiac Disease Prevalence by Leg-to-Trunk Fat Ratio Quartiles.

| Quartile | Ratio Range | Mean Ratio | N | Celiac Cases | Prevalence |
| --- | --- | --- | --- | --- | --- |
| Q1 (Lowest) | 0.22–0.69 | 0.584 | 708 | 3 | 0.42% |
| Q2 | 0.69–0.84 | 0.765 | 707 | 2 | 0.28% |
| Q3 | 0.84–1.02 | 0.926 | 707 | 4 | 0.57% |
| Q4 (Highest) | 1.02–2.55 | 1.228 | 708 | 2 | 0.28% |

*P for trend = 0.893 (Spearman correlation).*

### Sensitivity Analysis: Leanness Bias Control

To rule out the possibility that lower gynoid fat in celiac patients simply reflects overall leanness or malnutrition, we conducted sensitivity analyses stratified by BMI using survey-weighted estimates. First, we confirmed that BMI was similar between women with and without celiac disease (weighted mean 27.7 ± 8.0 vs. 29.2 ± 7.9 kg/m², p = 0.390), indicating that celiac patients in this population were not systematically underweight. Among the 11 celiac cases, none were underweight (BMI less than 18.5), four (36.4%) had normal weight, and seven (63.6%) were overweight or obese.

When the analysis was restricted to overweight and obese women only (BMI > 25 kg/m²), the association between celiac disease and lower gynoid fat persisted (**Supplementary Table S1**). Stratification by BMI category revealed that this divergence intensifies in obesity (**Supplementary Figure S2**), effectively ruling out malnutrition as a confounding factor. Among these clearly non-malnourished women, those with celiac disease (n = 7) had significantly lower weighted mean gynoid percent fat compared to those without celiac disease (40.6 ± 3.6% vs. 44.5 ± 4.6%, p = 0.005), representing an 8.7% relative reduction.

Stratification by BMI category revealed a consistent pattern across the weight spectrum using survey-weighted analyses. In normal weight women (BMI 18.5–25), celiac patients had 3.4% lower gynoid fat (weighted mean 38.1% vs. 39.4%, p = 0.229). In overweight women (BMI 25–30), celiac patients had 4.7% lower gynoid fat (weighted mean 41.2% vs. 43.2%, p = 0.024). The difference was most pronounced in obese women (BMI 30 or greater), where celiac patients had 11.3% lower gynoid fat (weighted mean 40.2 ± 4.6% vs. 45.3 ± 4.4%, p = 0.039) (Supplementary Table S2). These findings demonstrate that the association between celiac disease and lower gynoid fat is not explained by overall leanness or malnutrition and, if anything, becomes more pronounced with increasing body mass.

### Exploratory Immunometabolic Profile

To characterize the metabolic features of the lipedema phenotype, we compared immunometabolic markers between women with the lipedema phenotype (leg-to-trunk fat ratio above the 90th percentile, n = 283) and controls (n = 3,550). Women with the lipedema phenotype exhibited significantly lower systemic inflammation markers compared to controls. The neutrophil-to-lymphocyte ratio was 7.6% lower in women with the lipedema phenotype (1.96 ± 1.04 vs. 2.12 ± 1.03, p = 0.012), and total white blood cell count was 13.1% lower (6.47 ± 2.05 vs. 7.44 ± 2.31 × 10³/µL, p < 0.001).

Women with the lipedema phenotype also demonstrated markedly lower insulin resistance compared to controls. Median HOMA-IR was 44.2% lower in the lipedema phenotype group (1.35, IQR 0.95–2.01 vs. 2.42, IQR 1.47–4.30, p < 0.001) (Table 4, Figure 4). This favorable metabolic profile persisted across BMI categories. In normal weight women, median HOMA-IR was 8.7% lower in those with the lipedema phenotype (1.35 vs. 1.47, p = 0.116). The difference became more pronounced in overweight women, where median HOMA-IR was 34.7% lower (1.46 vs. 2.23, p = 0.002), and in women with class I obesity, where median HOMA-IR was 46.7% lower (1.76 vs. 3.31, p = 0.019). The magnitude of the metabolic advantage associated with the lipedema phenotype increased progressively with increasing BMI, suggesting that the metabolically favorable characteristics of gynoid fat distribution become more pronounced in overweight and obese women.

**Figure 4:**
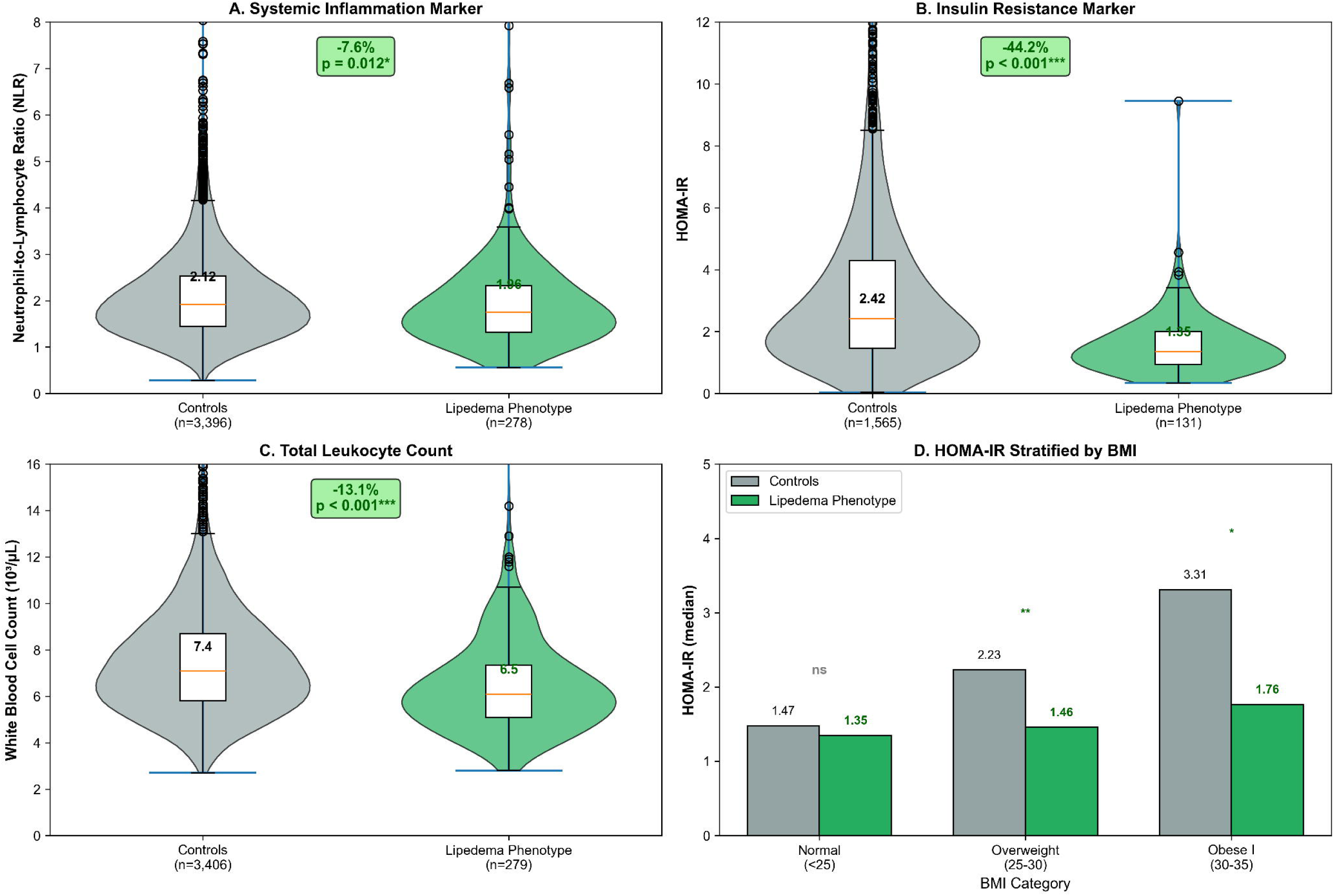
Exploratory Immunometabolic Profile: Lipedema Phenotype vs Controls. (A) Neutrophil-to-lymphocyte ratio (NLR), a marker of systemic inflammation, was significantly lower in women with the lipedema phenotype (p=0.012). (B) HOMA-IR, a marker of insulin resistance, was markedly lower in the lipedema phenotype group (p<0.001). (C) Total white blood cell count was significantly reduced in women with the lipedema phenotype (p<0.001). (D) HOMA-IR stratified by BMI category, demonstrating that the favorable metabolic profile persists across weight categories, with the largest differences observed in overweight and obese women. Green bars indicate the lipedema phenotype group; gray bars indicate controls. NHANES 2011–2014, adult women aged 20 years and older

**Table 4:**
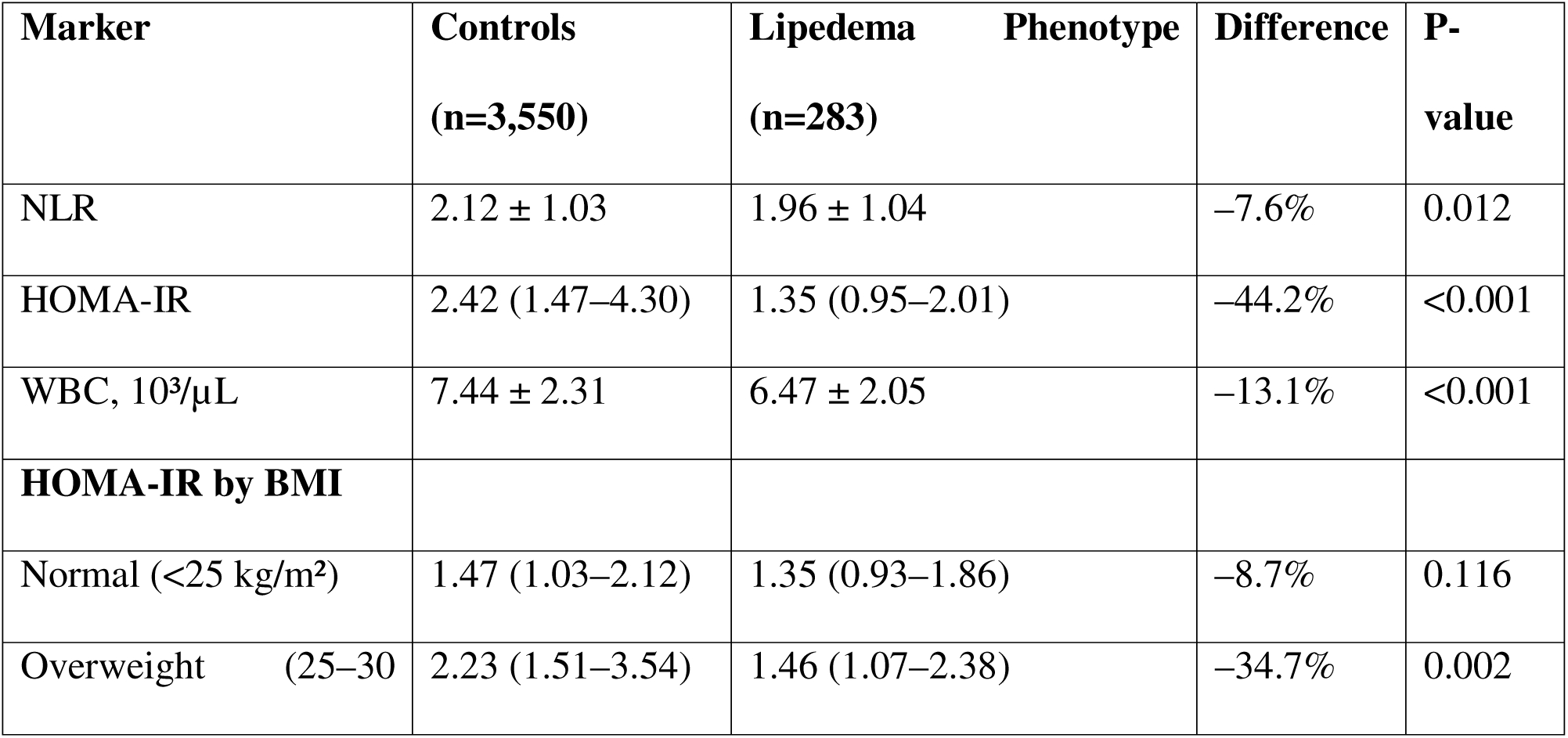
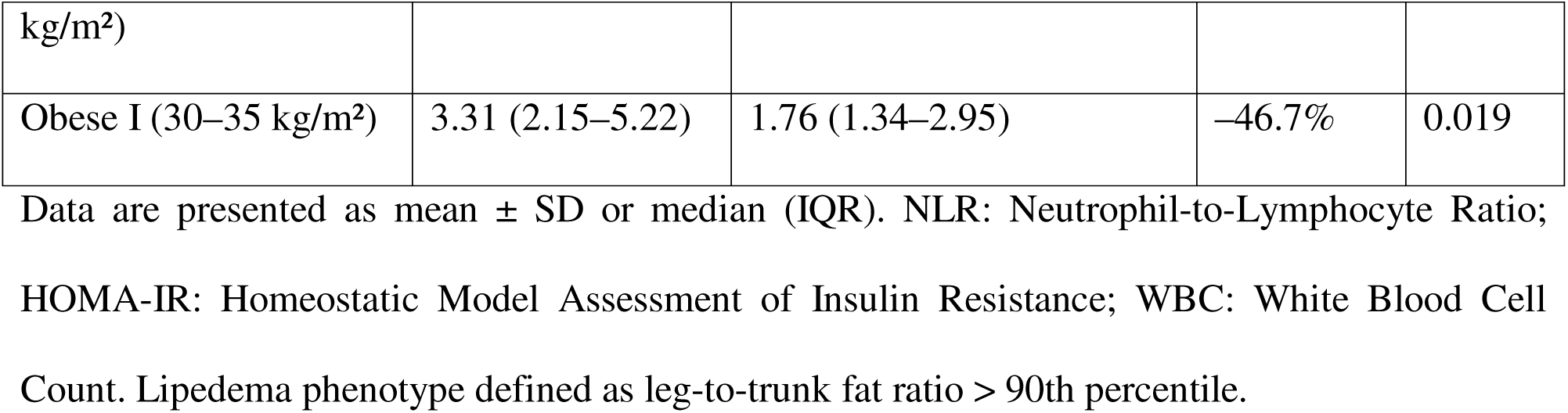
Immunometabolic Profile by Lipedema Phenotype Status.

## Discussion

This study presents the first epidemiological analysis contrasting the body composition of celiac disease against the lipedema-like phenotype in a nationally representative sample. Our findings reveal a significant phenotypic divergence: women with celiac disease exhibit reduced gynoid adiposity (p=0.0007), whereas the lipedema phenotype is characterized by a robust ’immunological shield’ profile—marked by significantly lower insulin resistance (HOMA-IR) and systemic inflammation (NLR). Although the strict serological definition of celiac disease limited the statistical power to detect a direct exclusion effect, the immunometabolic data strongly support the hypothesis that these conditions represent opposing physiological states: one of inflammatory catabolism and another of anabolic immune tolerance.

Unlike our initial hypothesis of a direct inverse association, the difference in lipedema phenotype prevalence between groups did not reach statistical significance (p=0.570). This is likely attributable to the strict ’gold-standard’ serological criteria used, which yielded a realistic but small number of active celiac cases (n=11, 0.56%), reducing statistical power (Type II error). **However, it is clinically relevant that complete exclusion was observed in the highest tier of gynoid adiposity, suggesting a potential threshold effect.** Furthermore, the significant reduction in gynoid fat percentage among celiac patients suggests that the **reduced abundance of this specific adipose depot is strictly associated with the disease**, contrasting sharply with the preservation seen in lipedema.

Furthermore, this immunometabolic distinction offers a mechanistic insight into the clinical utility of ketogenic diets for lipedema (21); the therapeutic benefit may stem not only from ketogenesis but also from the inherent exclusion of gluten, potentially mitigating inflammatory triggers in a phenotype that, while protected from overt celiac disease, remains sensitive to systemic inflammation.

### Biological Plausibility: Immunometabolic Antagonism

The most immediate interpretation of these findings aligns with the distinct metabolic profiles of regional adipose depots. Celiac disease is an autoimmune enteropathy driven by a Th1-dominant inflammatory response, characterized by elevated levels of interferon-gamma and TNF-alpha.

Conversely, the gluteofemoral adipose tissue characteristic of lipedema is metabolically distinct from the pro-inflammatory visceral fat associated with android obesity. Gluteofemoral fat acts as a "metabolic sink," efficiently trapping fatty acids and preventing ectopic lipid deposition, while secreting higher levels of adiponectin—an anti-inflammatory adipokine known to suppress Th1 differentiation and promote regulatory T-cell function. It is plausible that the massive expansion of this subcutaneous depot creates a systemic anti-inflammatory milieu that raises the threshold for autoimmune activation in genetically susceptible individuals, effectively antagonizing the pro-inflammatory state required for the phenotypic expression of celiac disease(22–26).

This ’immunological shield’ hypothesis is further supported by our exploratory metabolic analysis. We observed that women with the lipedema phenotype, even those with elevated BMI, displayed lower systemic inflammation (NLR) and **significantly** improved insulin sensitivity (HOMA-IR) compared to controls (**Table 4**, **Figure 4**). This aligns with the concept of ’Metabolically Healthy Obesity,’ where the expansion of subcutaneous gluteofemoral adipose tissue acts as a safe metabolic sink, sequestering lipids and preventing the systemic lipotoxicity and pro-inflammatory cytokine release that typically drive autoimmune dysregulation. The concept of the ’Immunological Shield’ is further supported by our secondary analysis of this cohort, which demonstrated that the lipedema phenotype is associated with lower odds against diabetes (OR 0.21, p < 0.001) and shows a significant dose-response inverse association with thyroid disease (**Supplementary Figure S3**). This confirms that the phenotypic definition used in this study captures a broadly protective immunometabolic profile, validating its potential to buffer against autoimmune dysregulation. Crucially, our sensitivity analysis reveals that this phenotypic divergence is not merely a consequence of malnutrition or the ’leanness bias’ often associated with celiac disease. In fact, the reduction in gynoid adiposity was most pronounced in the obese stratum (11.3% reduction, p=0.039) compared to normal-weight women. This suggests that even in a state of energy surplus, the celiac inflammatory milieu specifically antagonizes the expansion of the gluteofemoral ’metabolic sink’. This reinforces the concept that the lipedema phenotype and celiac autoimmunity represent biologically opposing states of adipose tissue function: one favoring indefinite subcutaneous expansion and immune tolerance, and the other promoting inflammation and restricting gynoid storage. Although adiponectin was not directly measured, the **substantial 44.2%** reduction in HOMA-IR observed in the lipedema phenotype serves as a robust physiological proxy. Extensive literature correlates high gluteofemoral adiposity with elevated adiponectin, which is the primary driver of the observed insulin sensitivity (27–30).

Crucially, this protection appears to extend beyond simple inflammation control to specific immune modulation. Recent findings regarding the "IgG Paradox" in lipedema strongly support this hypothesis. Previous clinical work by Amato et al.(31) demonstrated that while lipedema patients exhibit widespread food sensitivities (suggesting increased intestinal permeability), they paradoxically mount a blunted quantitative antibody response. This suggests a systemic downregulation of humoral immunity; essentially, the lipedema phenotype may maintain a state of "immune tolerance" that prevents the transition from gluten exposure to the overt autoimmune destruction characteristic of celiac disease, prioritizing energy conservation over immune hyper-vigilance.

Mechanistically, adiponectin likely plays a pivotal role in this suppression. As an abundant adipokine derived from the expanded subcutaneous tissue, adiponectin reduces the differentiation of CD4+ T cells into Th1 cells, decreasing the production of IFN-γ—the primary pro-inflammatory cytokine driving celiac pathology—in both obesity models and autoimmune conditions (32–35). This suppressive effect occurs through mechanisms involving the restriction of cellular glycolysis in Th1 cells—independent of the AMPK pathway—as well as the inhibition of co-stimulatory molecules such as CD40 and CD80 in antigen-presenting cells (33,35). While context-dependent pro-inflammatory effects have been described (36,37), the prevailing consensus points to a dominant immunosuppressive role of adiponectin regarding the Th1 response (32–35,38), reinforcing the biological plausibility of the lipedema phenotype as a buffer against celiac autoimmunity.

### The "Evolutionary Shield" Hypothesis

Extending beyond metabolism, our results support a alternative reinterpretation of lipedema through an evolutionary lens. We propose that the lipedema phenotype may represent an ancestral, adaptive mechanism for energy conservation and immunological stability rather than a primary pathology(14). In an evolutionary context, the ability to store energy safely in the lower extremities—without the metabolic penalties of visceral fat—would offer a survival advantage during reproductive years. The inverse association with celiac disease suggests that this "thrifty phenotype" may be coupled with a downregulated or distinct immune surveillance system. If lipedema represents a physiological state prioritized for tissue preservation and energy storage, it may naturally suppress the hyper-vigilant, destructive immune responses characteristic of autoimmunity. Thus, the disproportionate adipose tissue may function as an "immunological buffer" or shield, protecting the host from systemic autoimmune dysregulation(14,39–43).

The strongest evidence for the ’Immunological Shield’ comes from our exploratory analysis. The lipedema phenotype demonstrated a **44.2%** reduction in HOMA-IR and significantly lower NLR. Since high adiponectin levels are the primary driver of insulin sensitivity in gluteofemoral obesity , these HOMA-IR results serve as a powerful physiological proxy , confirming that the lipedema tissue is actively creating an anti-inflammatory, insulin-sensitizing milieu that is theoretically inhospitable to Th1-mediated autoimmunity.

### The Paradox of Genotype vs. Phenotype

These findings are particularly intriguing when juxtaposed with clinical genetic data. Previous studies have reported a high prevalence of HLA-DQ2 and HLA-DQ8 alleles—the primary genetic drivers of celiac disease—among women with lipedema(13). Our observation of reduced disease prevalence in this same phenotypic group suggests a decoupling of genotype and phenotype. The lipedema adipose tissue environment may exert a dominant epigenetic or immunomodulatory influence that prevents the clinical manifestation of celiac disease despite genetic susceptibility. This "penetrance suppression" reinforces the concept of the lipedema phenotype as a potent modifier of systemic immunity[13,46].

### Clinical Implications: A Call for Surgical Caution

The potential protective role of lipedema adipose tissue raises critical questions regarding current therapeutic paradigms, specifically the widespread use of large-volume liposuction. If the gluteofemoral depot functions as an active endocrine organ that suppresses autoimmunity, its aggressive removal could inadvertently disrupt this protective equilibrium[47]. The concept of "post-surgical adipose endocrine insufficiency" is already recognized in metabolic literature; our data suggest this risk may extend to the immune system. We hypothesize that the rapid removal of this immunomodulatory tissue could theoretically "unmask" latent autoimmune tendencies, leading to an increased incidence of conditions such as celiac disease or thyroiditis post-surgery. This possibility demands a shift in perspective: lipedema tissue should not be viewed merely as "diseased fat" to be excised, but as a functional organ whose systemic role must be understood before irreversible removal [6,8,16,17,26,48–52].

While our findings suggest a potential immunomodulatory role of gluteofemoral adipose tissue, prospective studies are essential before modifying current surgical approaches. We recommend establishing pre- and post-operative immunological monitoring protocols in lipedema surgery registries.

## Limitations

Our study has limitations inherent to its design. The cross-sectional nature of NHANES precludes causal inference, and the use of a DXA-derived proxy, while validated, cannot replace a clinical diagnosis of lipedema. We addressed the potential "leanness bias"—where celiac malabsorption prevents weight gain—by strictly stratifying our analysis to women with BMI ≥25 kg/m², ensuring we compared "overweight women with gynoid fat" to "overweight women with android fat." Furthermore, while we adjusted for ethnicity and smoking, residual confounding from unmeasured genetic or environmental factors remains possible. Limitations include the unavailability of specific inflammatory biomarkers such as hs-CRP and adiponectin in the 2011–2014 NHANES cycles, which prevented a direct assessment of the specific cytokine milieu.

## Future Directions

These findings necessitate a new direction in lipedema research. Longitudinal studies are urgently needed to track the long-term autoimmune health of women undergoing lipedema surgery. Registries should monitor not only pain and aesthetic outcomes but also the new onset of autoimmune diagnoses and markers of systemic inflammation. Mechanistic studies should profile the adipokine and cytokine secretome of lipedema tissue to directly test its suppressive effect on Th1 immune pathways. Until such data are available, the surgical management of lipedema should be approached with a nuanced understanding of the tissue’s potential systemic, protective functions. Future studies should directly quantify adiponectin levels to confirm if the expanded gluteofemoral depot in these patients retains its insulin-sensitizing properties, thereby counteracting Th1-mediated inflammatory pathways.

## Conclusion

In conclusion, this study identifies a robust **phenotypic divergence** between the lipedema phenotype and celiac disease in a high-risk population of women. These results challenge the view of lipedema solely as a burden, suggesting it may represent a distinct immunometabolic phenotype with inverse association against specific autoimmune conditions. This paradigm shift underscores the need for caution in surgical interventions and highlights the complexity of adipose tissue as an immunological organ.

## Data Availability

All data analyzed in this study are publicly available from the National Health and Nutrition Examination Survey (NHANES) at https://wwwn.cdc.gov/nchs/nhanes/. All analytic code used in this work is publicly available at https://github.com/alexandreamato/lipedema-celiac-nhanes.

https://wwwn.cdc.gov/nchs/nhanes/

https://github.com/alexandreamato/lipedema-celiac-nhanes

## Acknowledgements

ACMA, JLSA, and DAB designed research; ACMA conducted research and analyzed data; ACMA, JLSA, and DAB wrote the paper. ACMA had primary responsibility for final content. All authors have read and approved the final manuscript.

## Data Availability*

Data described in the manuscript are publicly available from the National Health and Nutrition Examination Survey (NHANES) website (https://wwwn.cdc.gov/nchs/nhanes/). The analytic code used for this study is publicly available at https://github.com/alexandreamato/lipedema-celiac-nhanes.git.

## Declaration of Generative AI and AI-assisted technologies in the writing process

During the preparation of this work the author(s) used Claude Opus 4.1 and GEMINI 3 Pro in order to assist in the generation and debugging of the Python code used for statistical analysis, specifically to ensure the correct implementation of NHANES complex survey design weights. After using this tool/service, the author(s) reviewed and edited the content as needed and take(s) full responsibility for the content of the published article.

BMI: Body Mass Index
CD: Celiac Disease
DXA: Dual-Energy X-ray Absorptiometry
EMA-IgA: Endomysial Antibody IgA
HOMA-IR: Homeostatic Model Assessment of Insulin Resistance
NHANES: National Health and Nutrition Examination Survey
NLR: Neutrophil-to-Lymphocyte Ratio
OR: Odds Ratio
Th1: T-helper 1
tTG-IgA: Tissue Transglutaminase IgA
WBC: White Blood Cell Count

---

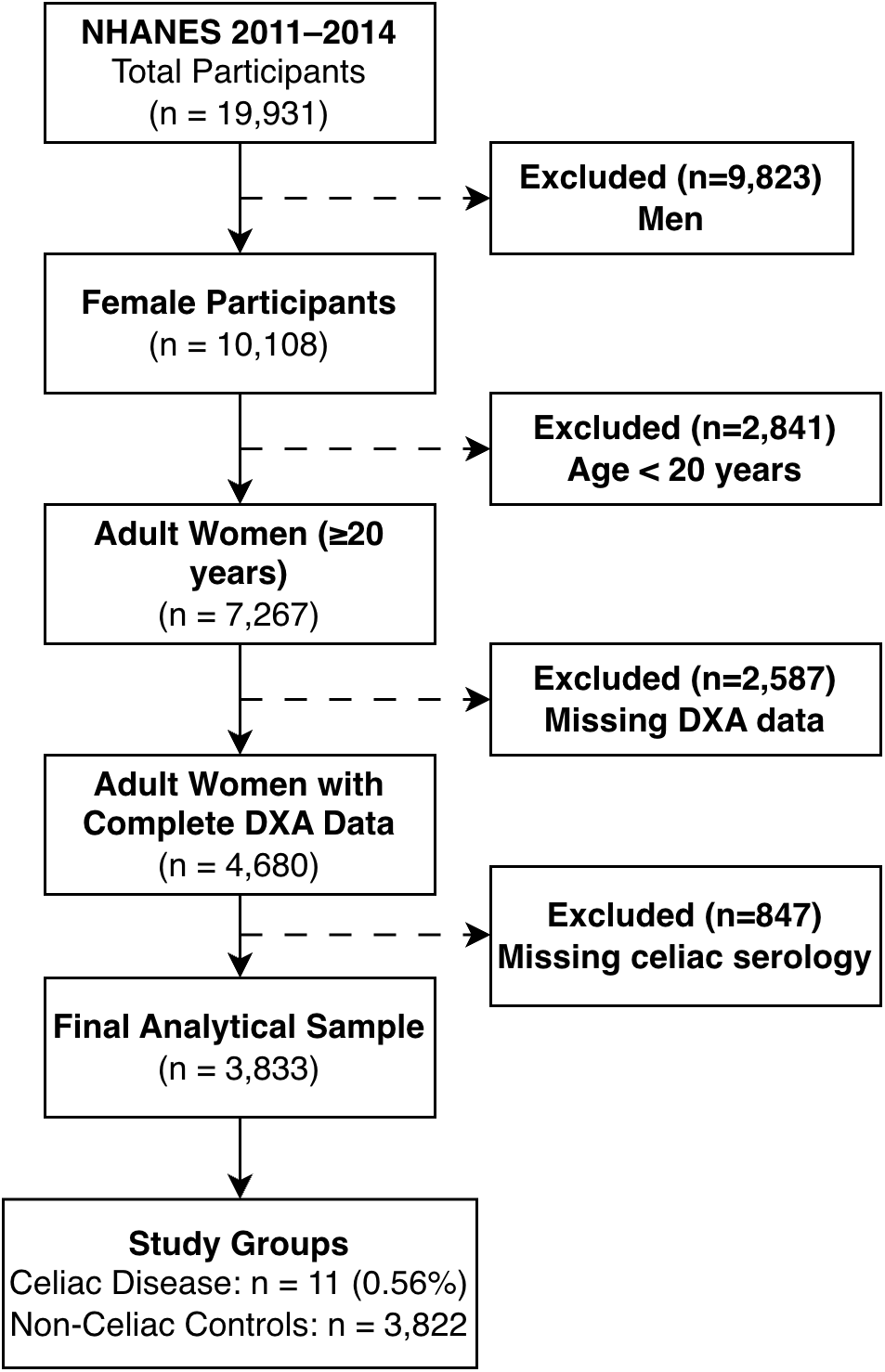

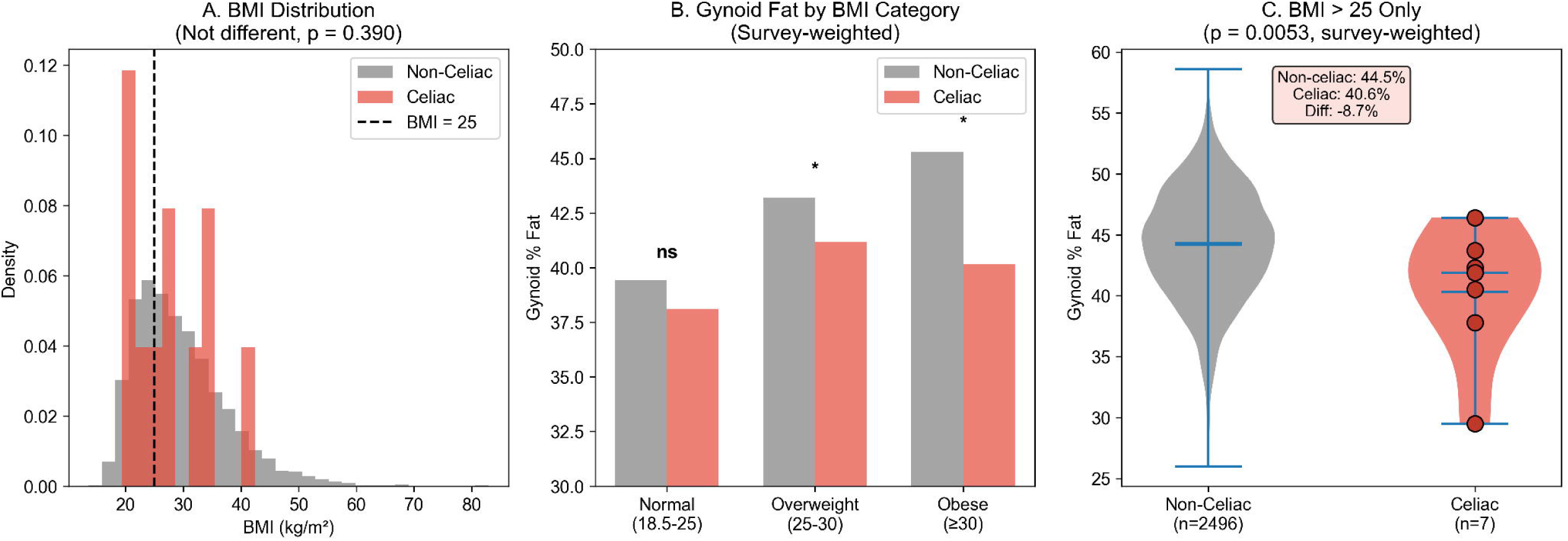

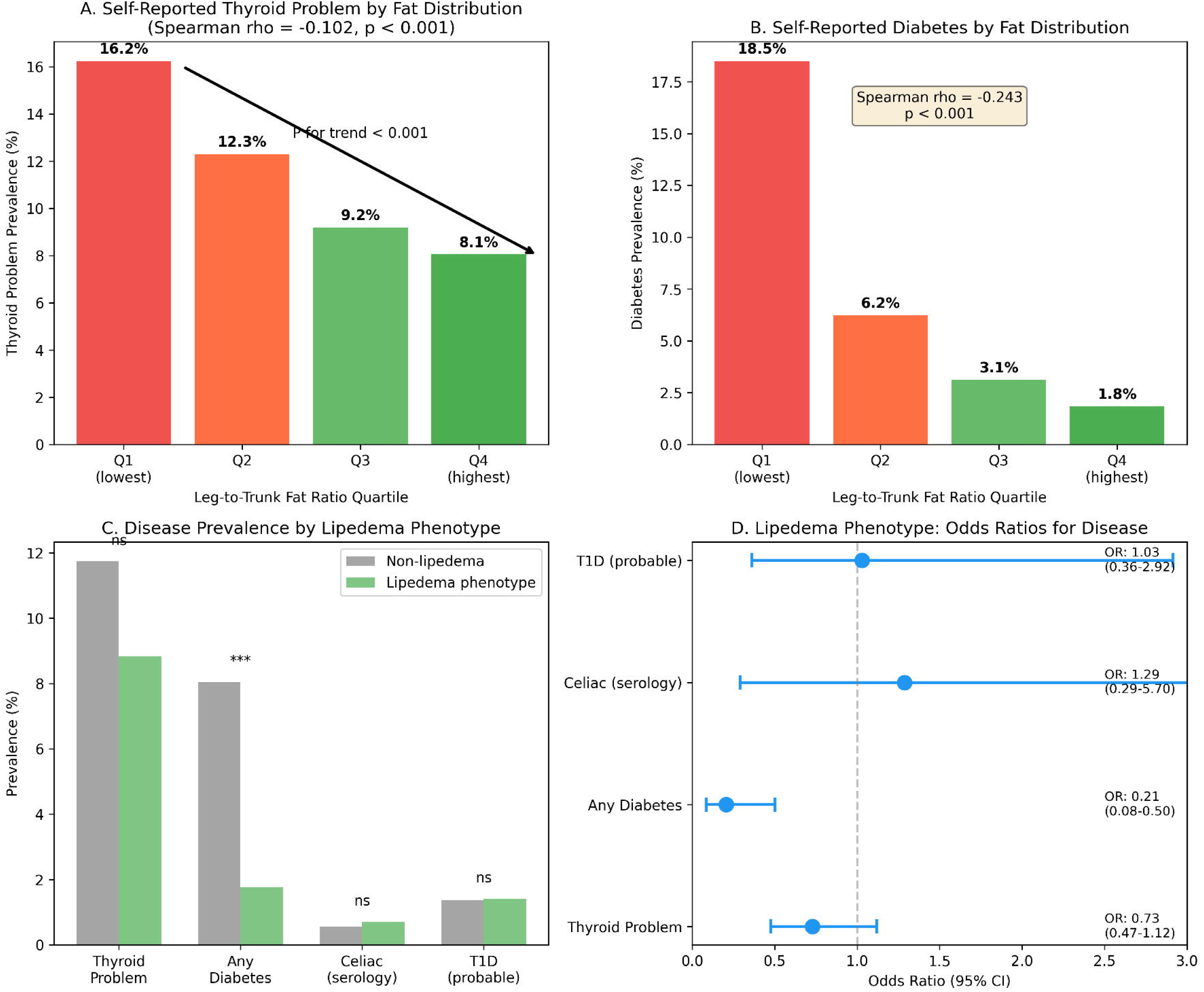

**supplementary Figure 1.**
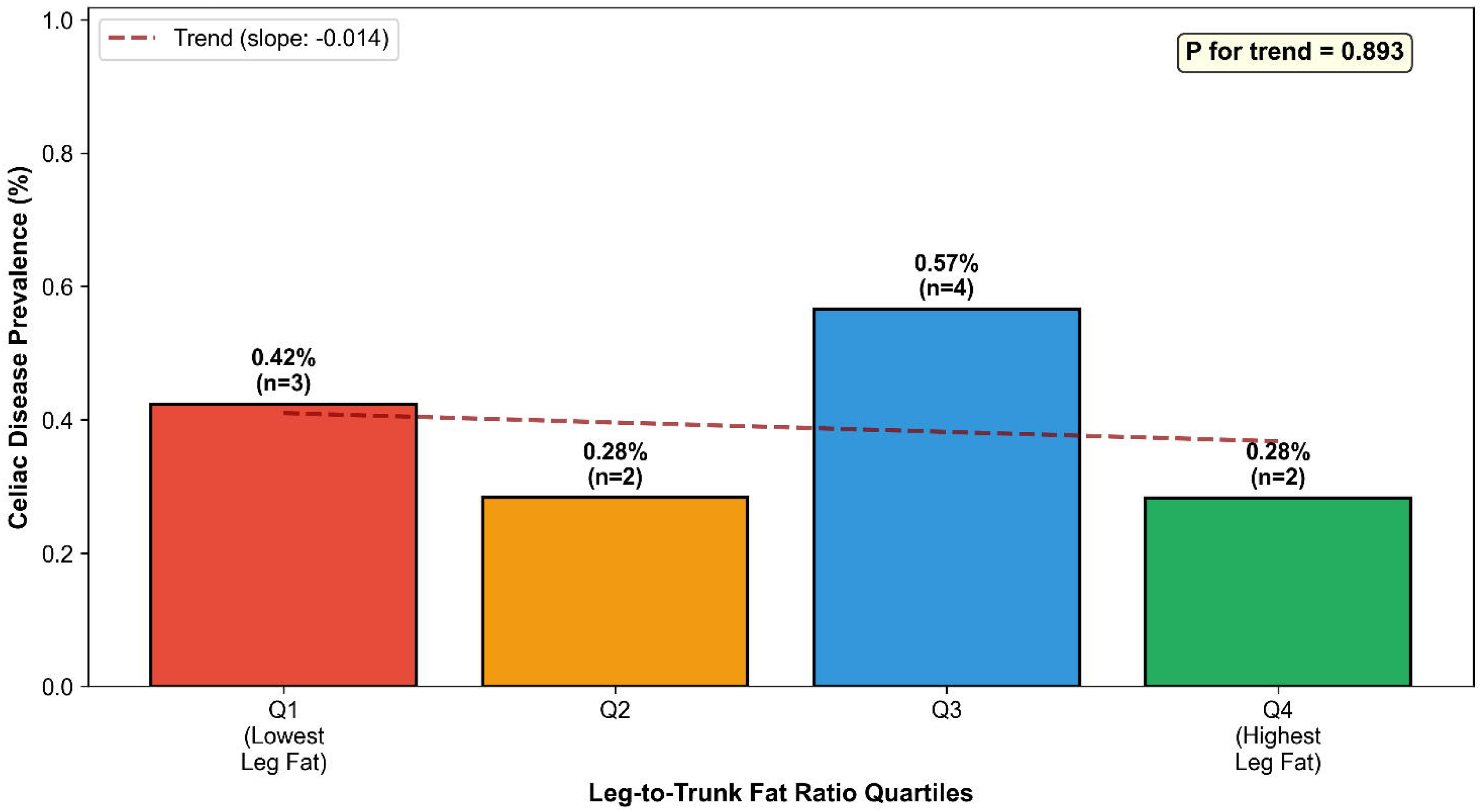
Celiac Disease Prevalence by Fat Distribution Quartiles NHANES 2011-2014, Adult Women (N=2,830)

## References

1. Amato ACM, Peclat APRM, Kikuchi R, Souza AC de, Silva MTB, Oliveira RHP de, Benitti DA, Oliveira JCP de. Brazilian Consensus Statement on Lipedema using the Delphi methodology. J Vasc Bras 2025;24.

2. Herbst KL, Kahn LA, Iker E, Ehrlich C, Wright T, McHutchison L, Schwartz J, Sleigh M, Donahue PMC, Lisson KH, et al. Standard of care for lipedema in the United States. Phlebology SAGE Publications Ltd; 2021;36:779–96.

3. Amato ACM, Amato JLS, Amato FCM, Benitti DA, Amato JLS, Benitti DA. Prevalência e fatores de risco para lipedema no Brasil. J Vasc Bras [Internet] 2022;21:1–11. Available from: http://www.scielo.br/scielo.php?script=sci_arttext&pid=S1677-54492022000100307&tlng=pt

4. Buso G, Depairon M, Tomson D, Raffoul W, Vettor R, Mazzolai L. Lipedema: A Call to Action! Obesity 2019;27:1567–76.

5. Straub LG, Funcke JB, Joffin N, Joung C, Al-Ghadban S, Zhao S, Zhu Q, Kruglikov IL, Zhu Y, Langlais PR, et al. Defining lipedema’s molecular hallmarks by multi-omics approach for disease prediction in women. Metabolism W.B. Saunders; 2025;168.

6. Grewal T, Kempa S, Buechler C. Lipedema: A Disease Triggered by M2 Polarized Macrophages? Biomedicines 2025;13:561.

7. Wolf S, Deuel JW, Hollmén M, Felmerer G, Kim BS, Vasella M, Grünherz L, Giovanoli P, Lindenblatt N, Gousopoulos E. A distinct cytokine profile and stromal vascular fraction metabolic status without significant changes in the lipid composition characterizes lipedema. Int J Mol Sci MDPI AG; 2021;22.

8. Kruppa P, Gohlke S, Łapiński K, Garcia-Carrizo F, Soultoukis GA, Infanger M, Schulz TJ, Ghods M. Lipedema stage affects adipocyte hypertrophy, subcutaneous adipose tissue inflammation and interstitial fibrosis. Front Immunol Frontiers Media SA; 2023;14.

9. Patton L, Ricolfi L, Bortolon M, Gabriele G, Zolesio P, Cione E, Cannataro R. Observational Study on a Large Italian Population with Lipedema: Biochemical and Hormonal Profile, Anatomical and Clinical Evaluation, Self-Reported History. Int J Mol Sci Multidisciplinary Digital Publishing Institute (MDPI); 2024;25.

10. Duhon BH, Phan TT, Taylor SL, Crescenzi RL, Rutkowski JM. Current Mechanistic Understandings of Lymphedema and Lipedema: Tales of Fluid, Fat, and Fibrosis. Int J Mol Sci 2022;23.

11. Felmerer G, Stylianaki A, Hollmén M, Ströbel P, Stepniewski A, Wang A, Frueh FS, Kim BS, Giovanoli P, Lindenblatt N, et al. Increased levels of VEGF-C and macrophage infiltration in lipedema patients without changes in lymphatic vascular morphology. Sci Rep Nature Research; 2020;10.

12. Kruglikov IL, Scherer PE. Is the endotoxin–complement cascade the major driver in lipedema? Trends in Endocrinology and Metabolism. Elsevier Inc.; 2024. p. 769–80.

13. Amato AC, Amato LL, Benitti D, Amato JL. Assessing the Prevalence of HLA-DQ2 and HLA-DQ8 in Lipedema Patients and the Potential Benefits of a Gluten-Free Diet. Cureus [Internet] 2023; Available from: https://www.cureus.com/articles/163916-assessing-the-prevalence-of-hla-dq2-and-hla-dq8-in-lipedema-patients-and-the-potential-benefits-of-a-gluten-free-diet

14. Amato AC. The Evolutionary Theory of Lipedema: A Perspective on Energy Storage and Chronic Inflammation. Cureus Springer Science and Business Media LLC; 2025;

15. Bouziat R, Hinterleitner R, Brown JJ, Stencel-Baerenwald JE, Ikizler M, Mayassi T, Meisel M, Kim SM, Discepolo V, Pruijssers AJ, et al. Reovirus infection triggers inflammatory responses to dietary antigens and development of celiac disease. Science (1979) 2017;356:44–50.

16. Wolf S, Rannikko JH, Virtakoivu R, Cinelli P, Felmerer G, Burger A, Giovanoli P, Detmar M, Lindenblatt N, Hollmén M, et al. A distinct M2 macrophage infiltrate and transcriptomic profile decisively influence adipocyte differentiation in lipedema. Front Immunol Frontiers Media S.A.; 2022;13.

17. Nankam PAN, Cornely M, Klöting N, Blüher M. Is subcutaneous adipose tissue expansion in people living with lipedema healthier and reflected by circulating parameters? Front Endocrinol (Lausanne) 2022;13.

18. Buso G, Favre L, Vionnet N, Gonzalez-Rodriguez E, Hans D, Puder JJ, Dubath C, Eap C Bin, Raffoul W, Collet TH, et al. Body Composition Assessment by Dual-Energy X-Ray Absorptiometry: A Useful Tool for the Diagnosis of Lipedema. Obes Facts S. Karger AG; 2022;15:762–73.

19. Dietzel R, Reisshauer A, Jahr S, Calafiore D, Armbrecht G. Body composition in lipoedema of the legs using dual-energy X-ray absorptiometry: A case-control study. British Journal of Dermatology 2015;173:594–6.

20. Nguyen VK, Middleton LYM, Huang L, Zhao N, Verly E, Kvasnicka J, Sagers L, Patel CJ, Colacino J, Jolliet O. Harmonized US National Health and Nutrition Examination Survey 1988-2018 for high throughput exposome-health discovery. 2023.

21. Amato ACM, Amato JLS, Benitti DA. The Efficacy of Ketogenic Diets (Low Carbohydrate; High Fat) as a Potential Nutritional Intervention for Lipedema: A Systematic Review and Meta-Analysis. Nutrients 2024;16.

22. Goossens GH. The Metabolic Phenotype in Obesity: Fat Mass, Body Fat Distribution, and Adipose Tissue Function. Obes Facts 2017;10:207–15.

23. Chait A, den Hartigh LJ. Adipose Tissue Distribution, Inflammation and Its Metabolic Consequences, Including Diabetes and Cardiovascular Disease. Front Cardiovasc Med 2020;7.

24. Booth AD, Magnuson AM, Fouts J, Wei Y, Wang D, Pagliassotti MJ, Foster MT. Subcutaneous adipose tissue accumulation protects systemic glucose tolerance and muscle metabolism. Adipocyte 2018;7:261–72.

25. De Fano M, Bartolini D, Tortoioli C, Vermigli C, Malara M, Galli F, Murdolo G. Adipose Tissue Plasticity in Response to Pathophysiological Cues: A Connecting Link between Obesity and Its Associated Comorbidities. Int J Mol Sci 2022;23:5511.

26. Gorgojo-Martínez JJ. Adipocentric Strategy for the Treatment of Type 2 Diabetes Mellitus. J Clin Med 2025;14:678.

27. Wang C, Mao X, Wang L, Liu M, Wetzel MD, Guan K-L, Dong LQ, Liu F. Adiponectin Sensitizes Insulin Signaling by Reducing p70 S6 Kinase-mediated Serine Phosphorylation of IRS-1. Journal of Biological Chemistry 2007;282:7991–6.

28. Borges MC, Oliveira IO, Freitas DF, Horta BL, Ong KK, Gigante DP, Barros AJD. Obesity-induced hypoadiponectinaemia: the opposite influences of central and peripheral fat compartments. Int J Epidemiol 2017;46:2044–55.

29. Lihn AS, Pedersen SB, Richelsen B. Adiponectin: action, regulation and association to insulin sensitivity. Obesity Reviews 2005;6:13–21.

30. Manolopoulos KN, Karpe F, Frayn KN. Gluteofemoral body fat as a determinant of metabolic health. Int J Obes 2010;34:949–59.

31. Amato AC, Amato JS, Benitti D. The IgG Paradox in Lipedema: More Food Sensitivities, Less Antibody Production. Cureus 2025;

32. Kim MW, Abid N bin, Jo MH, Jo MG, Yoon GH, Kim MO. Suppression of adiponectin receptor 1 promotes memory dysfunction and Alzheimer’s disease-like pathologies. Sci Rep 2017;7:12435.

33. Pham D-V, Park P-H. Adiponectin triggers breast cancer cell death via fatty acid metabolic reprogramming. Journal of Experimental & Clinical Cancer Research 2022;41:9.

34. Xiao Y, Deng T, Shang Z, Wang D. Adiponectin inhibits oxidization-induced differentiation of T helper cells through inhibiting costimulatory CD40 and CD80. Brazilian Journal of Medical and Biological Research 2017;50.

35. Surendar J, Frohberger SJ, Karunakaran I, Schmitt V, Stamminger W, Neumann A-L, Wilhelm C, Hoerauf A, Hübner MP. Adiponectin Limits IFN-γ and IL-17 Producing CD4 T Cells in Obesity by Restraining Cell Intrinsic Glycolysis. Front Immunol 2019;10.

36. Танянский ДА, Трулев АС, Агеева ЕВ, Никитин АА, Шавва ВС, Орлов СВ. Влияние адипонектина на продукцию аполипопротеинов А-1 и Е макрофагами человека. Молекулярная биология 2021;55:697–704.

37. Jung MY, Kim H-S, Hong H-J, Youn B-S, Kim TS. Adiponectin Induces Dendritic Cell Activation via PLCγ/JNK/NF-κB Pathways, Leading to Th1 and Th17 Polarization. The Journal of Immunology 2012;188:2592–601.

38. Pham D-V, Raut PK, Pandit M, Chang J-H, Katila N, Choi D-Y, Jeong J-H, Park P-H. Globular Adiponectin Inhibits Breast Cancer Cell Growth through Modulation of Inflammasome Activation: Critical Role of Sestrin2 and AMPK Signaling. Cancers (Basel) 2020;12:613.

39. Herbst KL. Rare adipose disorders (RADs) masquerading as obesity. Acta Pharmacol Sin [Internet] Nature Publishing Group; 2012;33:155–72. Available from: 10.1038/aps.2011.153

40. Klaus S. Adipose Tissue as a Regulator of Energy Balance. Curr Drug Targets 2004;5:241–50.

41. Child AH, Gordon KD, Sharpe P, Brice G, Ostergaard P, Jeffery S, Mortimer PS. Lipedema: An inherited condition. Am J Med Genet A [Internet] United States; 2010;152A:970–6. Available from: https://onlinelibrary.wiley.com/doi/10.1002/ajmg.a.33313

42. Grant RW, Dixit VD. Adipose tissue as an immunological organ. Obesity 2015;23:512–8.

43. Wells JCK. The evolution of human fatness and susceptibility to obesity: an ethological approach. Biological Reviews 2006;81:183–205.

44. Atz JW. Effects of hybridization on pigmentation in fishes of the genus Xiphophorus. Zool Sci Contrib N Y Zool Soc 1962;47:153–81.

45. Kenny PA, Bissell MJ. Tumor reversion: Correction of malignant behavior by microenvironmental cues. Int J Cancer 2003;107:688–95.

46. Amato AC, Amato JL, Benitti D. Efficacy of Liposuction in the Treatment of Lipedema: A Meta-Analysis. Cureus [Internet] 2024; Available from: https://www.cureus.com/articles/232839-efficacy-of-liposuction-in-the-treatment-of-lipedema-a-meta-analysis

47. Pinnick KE, Nicholson G, Manolopoulos KN, McQuaid SE, Valet P, Frayn KN, Denton N, Min JL, Zondervan KT, Fleckner J, et al. Distinct developmental profile of lower-body adipose tissue defines resistance against obesity-associated metabolic complications. Diabetes 2014;63:3785–97.

48. Camastra S, Vitali A, Anselmino M, Gastaldelli A, Bellini R, Berta R, Severi I, Baldi S, Astiarraga B, Barbatelli G, et al. Muscle and adipose tissue morphology, insulin sensitivity and beta-cell function in diabetic and nondiabetic obese patients: Effects of bariatric surgery. Sci Rep Nature Publishing Group; 2017;7.

49. Kane H, Lynch L. Innate Immune Control of Adipose Tissue Homeostasis. Trends Immunol 2019;40:857–72.

50. Karpe F, Pinnick KE. Biology of upper-body and lower-body adipose tissue—link to whole-body phenotypes. Nat Rev Endocrinol 2015;11:90–100.

51. Lopategi A, López-Vicario C, Alcaraz-Quiles J, García-Alonso V, Rius B, Titos E, Clària J. Role of bioactive lipid mediators in obese adipose tissue inflammation and endocrine dysfunction. Mol Cell Endocrinol 2016;419:44–59.

