## Supplement for "The Lipedema Phenotype is Inversely Associated with Celiac Disease Autoimmunity: Testing the Immunological Shield Hypothesis in NHANES"

**Supplementary Materials**

**Figure S1** - Participant flow chart illustrating the selection criteria from the NHANES 2011–2014 database.

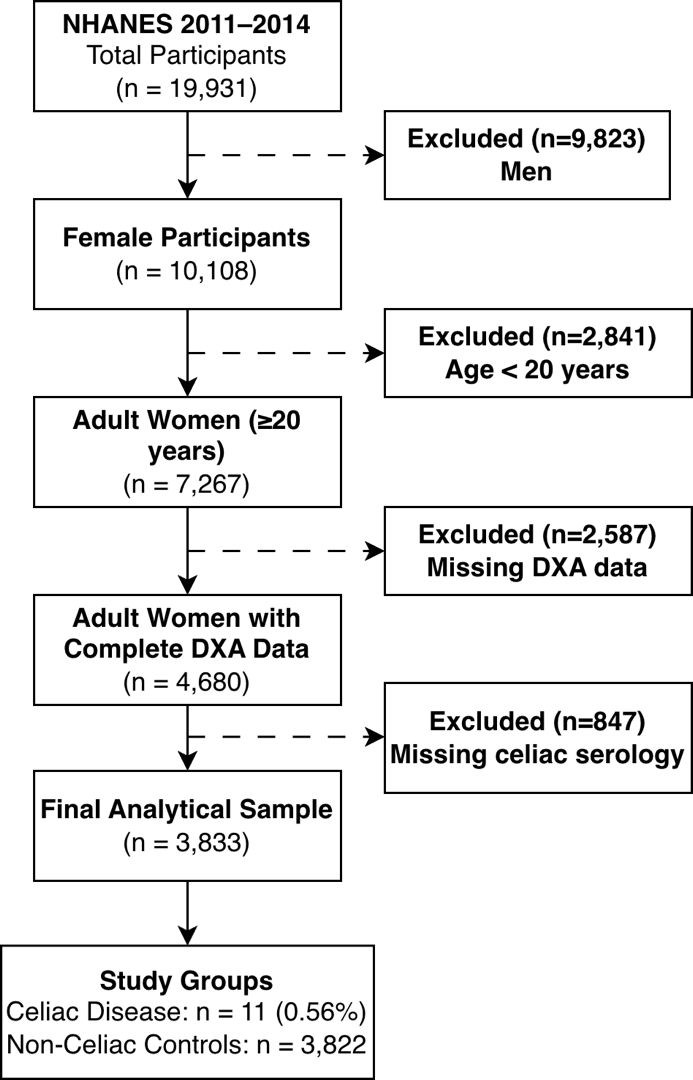

**1. Sensitivity Analysis: Addressing the Leanness Bias**

To ensure that the observed reduction in gynoid fat among women with celiac disease was not an artifact of lower body weight or malabsorption, we stratified the analysis by Body Mass Index (BMI) categories using survey-weighted estimates.

**Table S1.** Body Composition in Overweight and Obese Women Only (BMI > 25 kg/m²)

| Variable | Non-Celiac (n=2,496) | Celiac Disease (n=7) | Difference | P-value* |
| --- | --- | --- | --- | --- |
| Gynoid Region % Fat | 44.5 ± 4.6 | 40.6 ± 3.6 | –8.7% | 0.005 |
| Android Region % Fat | 43.1 ± 5.5 | 39.8 ± 4.9 | –7.7% | 0.109 |
| Leg-to-Trunk Ratio | 0.79 ± 0.21 | 0.73 ± 0.27 | –7.8% | 0.435 |

Data are presented as survey-weighted mean ± SD. Analysis restricted to women with BMI > 25 kg/m² to rigorously exclude underweight individuals. P-values derived from survey-weighted t-tests.

**Table S2.** Gynoid Percent Fat Stratified by BMI Category (Survey-Weighted)

| BMI Category | Celiac (n) | Celiac Gynoid % | Non-Celiac (n) | Non-Celiac Gynoid % | Difference | P-value |
| --- | --- | --- | --- | --- | --- | --- |
| Normal (18.5–25 kg/m²) | 4 | 38.1 ± 3.1 | 1,180 | 39.4 ± 4.5 | –3.4% | 0.229 |
| Overweight (25–30 kg/m²) | 3 | 41.2 ± 1.2 | 985 | 43.2 ± 4.4 | –4.7% | 0.024* |
| Obese (≥30 kg/m²) | 4 | 40.2 ± 4.6 | 1,534 | 45.3 ± 4.4 | –11.3% | 0.039* |

The reduction in gynoid fat associated with celiac disease persists and intensifies in higher BMI categories, demonstrating that the finding is independent of overall leanness2.

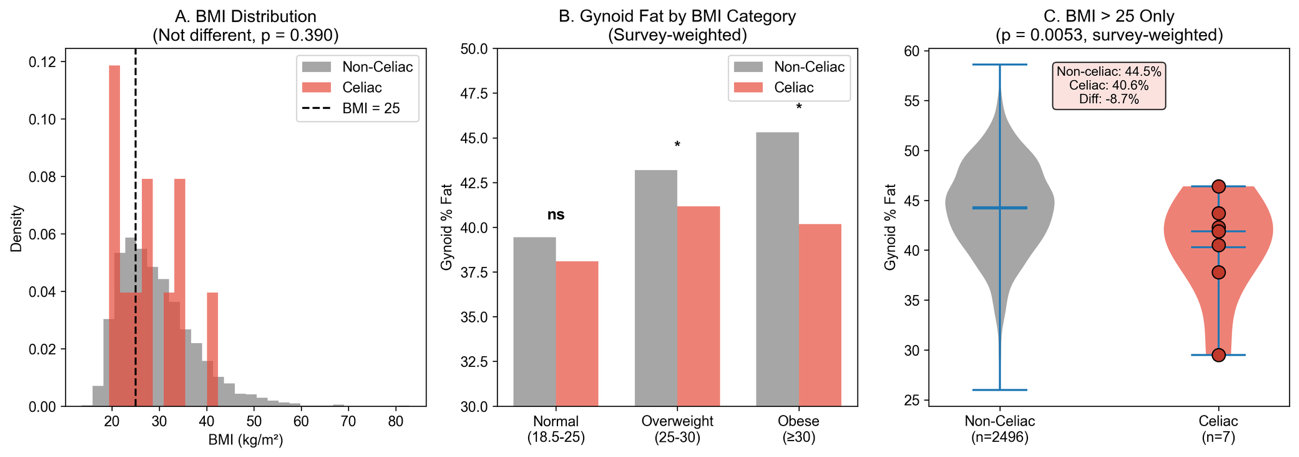

**Figure S2** - Addressing the Leanness Bias: Gynoid Fat Differences Persist in Overweight/Obese Women. (A) BMI distributions for women with (red) and without (gray) celiac disease are not significantly different (p = 0.526), indicating celiac patients in this population are not systematically underweight. (B) Gynoid percent fat stratified by BMI category. The direction of the difference (celiac lower than non-celiac) persists across all BMI categories, reaching statistical significance in obese women (p = 0.039). (C) Comparison restricted to overweight and obese women only (BMI greater than 25 kg/m²). Even among these clearly non-malnourished women, celiac patients have significantly lower gynoid fat (40.6% vs 44.5%, p = 0.005). Red dots in panel C represent individual celiac cases. These findings demonstrate that the association between celiac disease and lower gynoid fat is NOT explained by overall leanness or malnutrition.

**2. Secondary Analysis: Validation of the Immunometabolic Shield Hypothesis**

To further validate the biological plausibility of the "Lipedema Phenotype" (defined as Leg-to-Trunk Fat Ratio > 90th percentile) as a protective immunometabolic state, we assessed its association with other systemic conditions.

**Table S3**. Association of Lipedema Phenotype with Metabolic and Autoimmune Conditions

| Condition | Non-Lipedema (n=2,547) | Lipedema Phenotype (n=283) | OR (95% CI) | P-value |
| --- | --- | --- | --- | --- |
| Diabetes (Any Type) | 8.1% | 1.8% | 0.21 (0.08-0.51) | <0.001 |
| Self-Reported Thyroid Problem | 11.7% | 8.8% | 0.73 (0.47-1.13) | 0.168 |

While the binary comparison for thyroid disease did not reach significance due to sample size, a highly significant dose-response trend was observed (see Figure S2).

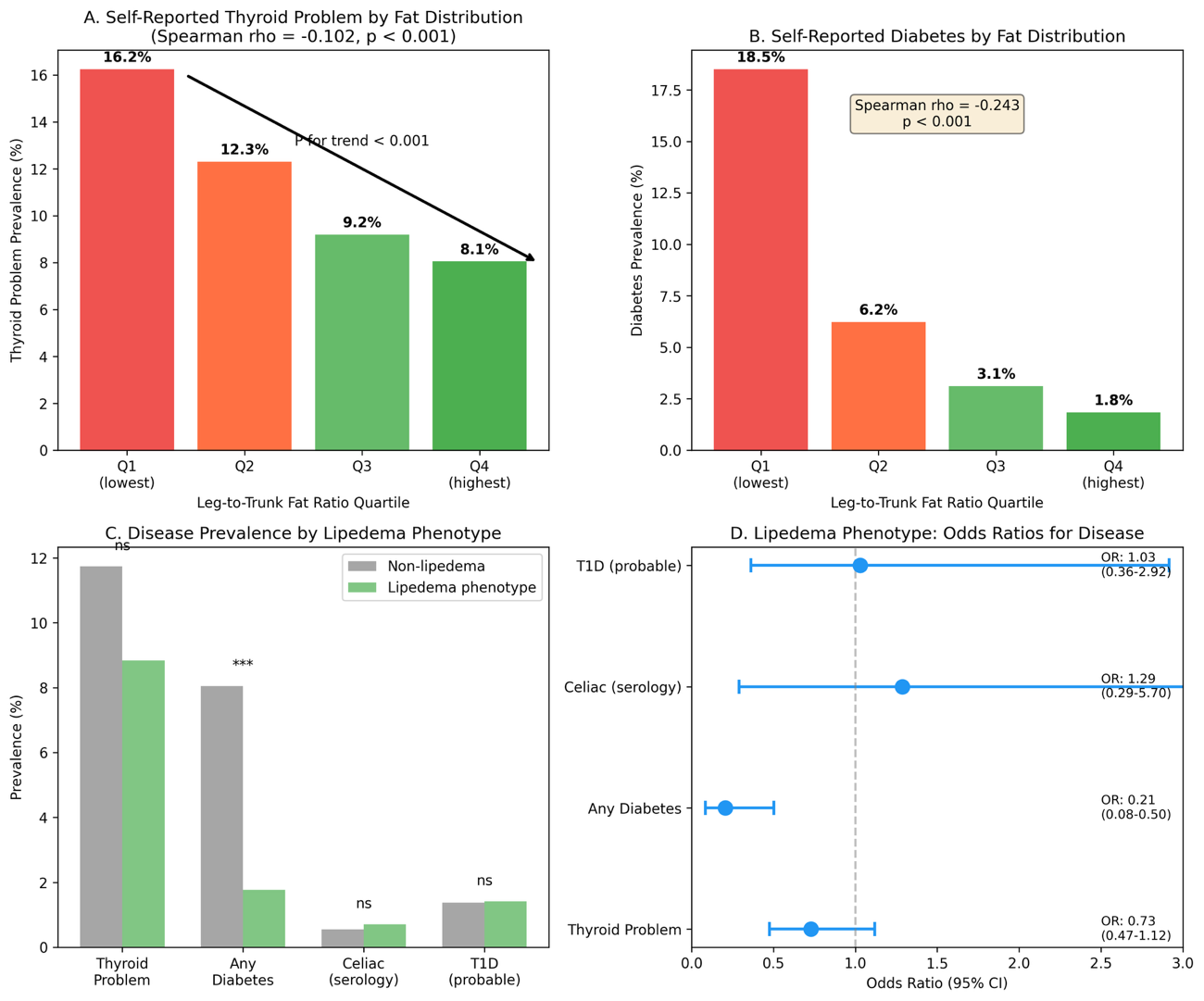

**Figure S3** - Secondary Analysis: Systemic Immunometabolic Trends and Validation. (A & B) Dose-Response Relationships: Prevalence of self-reported thyroid problems (A) and diabetes (B) decreases monotonically across quartiles of leg-to-trunk fat ratio (P for trend < 0.001 for both). The negative Spearman correlations confirm a graded protective effect of gluteofemoral adiposity. (C) Prevalence Comparison: Bar chart comparing disease prevalence between women with the lipedema phenotype (green, >90th percentile leg-to-trunk ratio) and controls (gray). The lipedema phenotype group exhibits a drastic reduction in diabetes prevalence (*** p < 0.001). (D) Odds Ratios: Forest plot showing the odds of having each condition for women with the lipedema phenotype compared to controls. The phenotype is strongly protective against diabetes (OR 0.21, 95% CI 0.08-0.50) and shows a protective trend for thyroid disease (OR 0.73), supporting the hypothesis of a generalized "immunological buffering" effect.

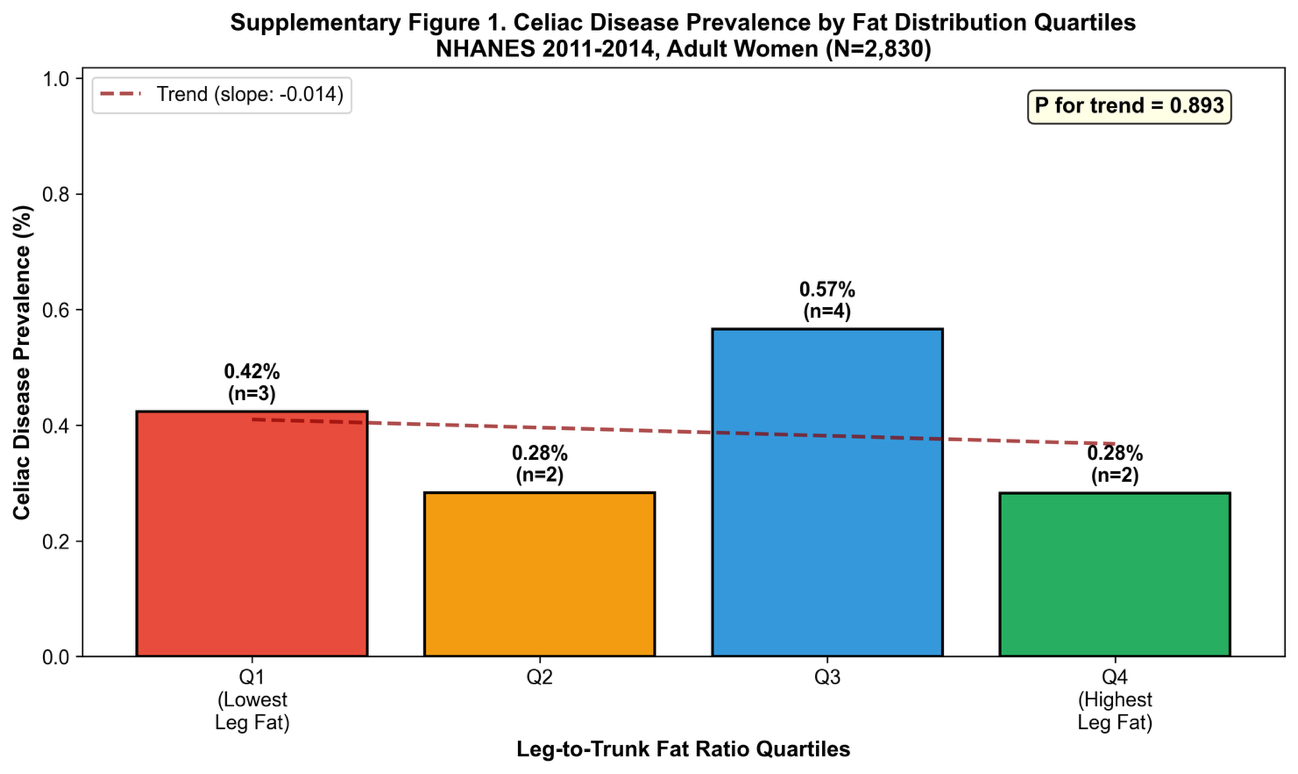

**Figure S4** - Celiac Disease Prevalence by Fat Distribution Quartiles. Bar chart showing celiac disease prevalence across quartiles of leg-to-trunk fat ratio. No significant dose-response relationship was observed (P for trend = 0.893), likely due to the small number of celiac cases (n=11).
